# Prevalence and predictors of depression, anxiety and stress symptoms among pregnant women during COVID-19-related lockdown in Abakaliki, Nigeria

**DOI:** 10.1101/2020.08.30.20184697

**Authors:** Johnbosco I. Nwafor, Ijeoma N. Okedo-Alex, Arinze C. Ikeotuonye

**Author notes:** **Corresponding author:** Johnbosco Ifunanya Nwafor, Department of Obstetrics and Gynaecology, Alex Ekwueme Federal University Teaching Hospital Abakaliki, Nigeria. Email.

## Abstract

**Introduction:** Several studies on COVID-19 and pregnancy have been published recently, but few studies have evaluated the impact of this pandemic on maternal mental health particularly in low-resource setting.

**Aim:** To determine the prevalence and predictors of COVID-19-related depression, anxiety and stress symptoms among pregnant women.

**Materials and methods:** This was a questionnaire-based cross-sectional study conducted among 456 pregnant women attending prenatal care at Abakaliki, Nigeria during COVID-19-related lockdown. They were screened for psychological morbidities using DASS 21 (Depression, Anxiety and Stress Scale).

**Results:** Severe and extremely severe depression were reported in 33 (7.2%) and 29 (6.4%) participants respectively. 15 (3.3%) and 35 (7.7%) women had severe and extremely severe anxiety respectively. 105 (23%) had severe anxiety whereas 76 (16.7%) reported extremely severe stress. Multiparity (2 - 4) and occupations such as trading and farming were predictors of depression whereas grandmultiparity, urban residence and trading were identified as predictors of anxiety and stress.

**Conclusion:** Depression, anxiety and stress symptoms were relatively common among pregnant women during COVID-19-related lockdown in Abakaliki, Nigeria. There is a need to integrate screening for depression, anxiety and stress in existing antenatal care programs so as to identify and prevent long term adverse psychological outcome related to COVID-19 pandemic.

## INTRODUCTION

In December, 2019, a series of pneumonia cases caused by SARS-CoV-2 emerged in Wuhan, China.^1^ The infection has spread to over 110 countries including Nigeria prompting World Health Organization to declare it a pandemic on March 11, 2020.^1^ Nigeria Centre for Disease Control announced the first confirmed case of coronavirus disease in Nigeria on February 27, 2020 and since then many confirmed cases have been reported in all the States across the country.^2^ By August 24, 2020, there were 52,548 confirmed cases and 1,004 deaths from SARS-CoV-2 infection in Nigeria.^2^ Globally, about 10% of pregnant women suffer from mental disorder in normal times, primarily depression^3^ and it has been reported to be higher (16%) in developing nations^4^. This may be aggravated during COVID-19 pandemic when pregnant women may have restricted access to mental health services. The mental health impact of the COVID-19 pandemic on childbearing women is a major public health challenge, which require appropriate and timely health care support to avert adverse health outcomes.

The information on the possible long term physical and psychological impact of the coronavirus on pregnant women and the foetus is still not clear. Studies on the psychological effects of the SARS outbreak in 2003 on the experience of pregnant women show that, in addition to the physical health implications, they experienced high levels of depression, anxiety and stress about being infected.^5^ Anxiety over the pregnant woman’s own health and that of the foetus may have detrimental effects on maternal and foetal well-being.^6^

Anxiety is common during and after pregnancy as women anticipate and adjust to motherhood, especially in those women and couples who have previously experienced adverse pregnancy outcomes, such as, miscarriage and perinatal death.^7,8^ The COVID-19 pandemic has resulted to fear, anxiety and concern in many nations as a result of the pandemic itself, but also due to the restrictive public health measures implemented to reduce community transmission. Border closures, travel bans, quarantine measures, physical distancing have resulted in increased isolation and decreased access to, and interaction with, social supports and networks. This is likely to result in increased stress, anxiety, loneliness and depression, particularly for pregnant women who will have an added level of concern about their own health and protecting their unborn baby. Empirical evidence suggests that prenatal stress is related to higher rates of adverse birth outcomes, such as preterm delivery, low birth weight and high rate of caesarean section.^7,8^

Measures and guidelines were altered in the management of clients including pregnant women in order to prevent transmission of the virus. Pregnant women are likely to be affected by the significant changes in the management of pregnancy, labour, birth and postnatal care provided by health services including a reduction in face-to-face appointments and use of telehealth.^8^ Studies have shown that lockdown is creating significant stress on pregnant women with flow on effects to the maternity providers (obstetricians, midwives, nurses, allied health professionals) caring for them.^9^ In Nigeria, the psychological impact of these measures on the pregnant women is yet to be described. Therefore, the objectives of this study was to determine the prevalence and associated factors of depression, anxiety and stress among pregnant women attending prenatal care in a tertiary health institution in Abakaliki during COVID-19-related lockdown in Nigeria.

## MATERIALS AND METHOD

### Study design, period and setting

This was a questionnaire-based cross-sectional study conducted from March 1, 2020 to July 31, 2020 at the antenatal clinic of Alex Ekwueme Federal University Teaching Hospital, Abakaliki, Nigeria. The hospital manages about 4500 deliveries annually and receives referral from all parts of the state and neighboring states of Benue, Enugu, Cross River and Abia as well as any part of the country.^10^

### Study participants and criteria

The study participants were pregnant women attending prenatal care at the study facility during the study period. Women who gave consent to participate in the study were included. Those with history of psychiatric illness or conditions that limit their ability to understand the study questions were excluded from the study.

### Sampling technique

A total sampling method was used in the study and consecutive pregnant women who were attending antenatal care were approached to participate in the study.

### Sample size determination

Considering the prevalence (p) of depression among pregnant women to be 13.5% based on study by Taubman-Ben-Ari et al.,^6^ the sample size was 208 at precision of 5% and design effect of 1.5 using OpenEpi software.^11^ If estimated considering the prevalence of stress (p=29.8%) and anxiety (p=24.1%), the sample size, with the other assumptions remaining constant, would be 315 and 288 respectively. Considering the largest value among the three, 315 and assuming an approximate 10% of questionnaires to have incomplete responses, the sample size was estimated to be 346.5. However, 456 pregnant women participated in the study.

### Data collection instrument

The data was collected using self-administered structured questionnaire. The questionnaire had two components that included socio-demographic characteristics and Depression Anxiety and Stress Scale-21 (DASS-21).

The sociodemographic variables collected were age, parity, marital status, area of residence, educational level attainment, occupation and semester of gestation.

DASS-21 is a self-reporting, 21-item questionnaire where seven questions each elicit information regarding depression, anxiety and stress subscales.^12^ In completing the DASS-21, the individual is required to indicate the presence of a symptom over the previous week. Each item was scored from 0 (“did not apply to me at all over the past week”) to 3 (“applied to me very much or most of the time over the past week”). In this tool, a 4-point severity scale is used for assessing the state of the participants in the three subscales over the past week. Each score is multiplied by 2, and the final scores were assessed against the DASS severity ratings [Table 1].

**Table 1:** Depression Anxiety and Stress Scale severity ratings

| Severity | Depression | Anxiety | Stress |
| --- | --- | --- | --- |
| Normal | 0 - 9 | 0 - 7 | 0 - 14 |
| Mild | 10 - 13 | 8 - 9 | 15 - 18 |
| Moderate | 14 - 20 | 10 - 14 | 19 - 25 |
| Severe | 21 - 27 | 15 - 19 | 26 - 33 |
| Extremely severe | ≥ 28 | ≥ 20 | ≥ 34 |

The survey questionnaire and the consent form in English, were translated into Igbo language, and then back-translated to English to check for semantic equivalence. The reliability of the questionnaire was checked by conducting a pretest among pregnant women in the antenatal clinic, by taking 5% of the sample size. From the pretest, understandability, clarity, and organization of the questionnaire were checked. From the reliability test, 0.896 Cronbach’s alpha value was found.

### Ethical considerations

This study was approved by the Research and Ethics Committee of the Alex Ekwueme Federal University Teaching Hospital, Abakaliki. Informed consent was taken from the study participants after informing the study subjects on study objectives, expected outcomes, and benefits associated with it. Confidentiality of responses was maintained throughout the study.

### Data processing and analysis

All returned questionnaires were checked manually for completeness and consistency of responses. The collected data were entered and analyzed using SPSS version 22 (IBM Corp. Amork, New York, U.S.A). Continuous variables were presented as means ± standard deviations (SDs), while categorical variables were summarized as numbers and percentages. Multivariate logistic regression analysis, presented with the odds ratios (OR) and 95% confidence intervals (CI), was used to identify socio-demographic factors associated with psychological morbidities. The P values < 0.05 were considered to be statistically significant.

## RESULTS

456 women participated in the study. The mean age of the participants was 27 ± 12.6, ranged from 18 to 45 years and the modal age was 28 −37 years. Majority of the participants were married (87.9%), para 2-4 (36.2%), resided in urban area (61.2%), had secondary education (49.1%), were traders (38.4%) and were in 2^nd^ trimester gestation (45.8%) [Table 2].

**Table 2:** Socio-demographic characteristics of the participants (N = 456)

| Variable | Frequency (%) |
| --- | --- |
| Age, yr |  |
| 18 - 27 | 160 (35.1) |
| 28 - 37 | 197 (43.2) |
| 38 - 45 | 99 (21.7) |
| Mean age, yr ± SD | |
| Parity |  |
| 0 | 95 (20.8) |
| 1 | 107 (23.5) |
| 2 - 4 | 165 (36.2) |
| $\geq 5$ | 89 (19.5) |
| Marital status |  |
| Single | 25 (5.5) |
| Married | 401 (87.9) |
| Divorced | 13 (2.9) |
| Widowed | 17 (3.7) |
| Area of residence |  |
| Rural | 177 (38.8) |
| Urban | 279 (61.2) |
| Educational qualification |  |
| No formal education | 31 (6.8) |
| Primary | 66 (14.5) |
| Secondary | 224 (49.1) |
| Tertiary | 135 (29.6) |
| Occupation |  |
| House wife | 81 (17.8) |
| Farmer | 101 (22.1) |
| Trader | 175 (38.4) |
| Artisan | 36 (7.9) |
| Civil servant | 63 (13.8) |
| Semester of gestation |  |
| 1<sup>st</sup> trimester | 88 (19.3) |
| 2<sup>nd</sup> trimester | 209 (45.8) |
| 3<sup>rd</sup> Trimester | 159 (34.9) |

Table 3 shows the prevalence of various psychological state of the study cohorts. Following DAS 21 assessment, depression was reported in 206 (45.2%) participants. 144 (31.6%) had mild-to-moderate depression, 33 (7.2%) had severe depression and 29 (6.4%) had extremely severe depression. 171 (37.5%) women reported anxiety symptoms. Of these, mild-to-moderate anxiety occurred in 121 (26.5%) participants. Severe and extremely severe anxiety were seen in 15 (3.3%) and 35 (7.7%) of the study cohorts respectively. Of 456 participants, 259 (56.8%) had stress symptoms. Seventy-eight (17.1%) women were found to have mild-to-moderate stress symptoms, whereas 105 (23%) and 76 (16.7%) reported severe and extremely severe stress respectively.

**Table 3:** Psychological outcome using DAS 21 among the participants (N = 456)

| Psychological category | Frequency (%) |
| --- | --- |
| Depression |  |
| Normal | 250 (54.8) |
| Mild | 83 (18.2) |
| Moderate | 61 (13.4) |
| Severe | 33 (7.2) |
| Extremely severe | 29 (6.4) |
| Anxiety |  |
| Normal | 285 (62.5) |
| Mild | 67 (14.7) |
| Moderate | 54 (11.8) |
| Severe | 15 (3.3) |
| Extremely severe | 35 (7.7) |
| Stress |  |
| Normal | 197 (43.2) |
| Mild | 25 (5.5) |
| Moderate | 53 (11.6) |
| Severe | 105 (23.0) |
| Extremely severe | 76 (16.7) |

Multivariable logistic regression analysis showing the predictors of depression, anxiety and stress among the participants is shown in Table 4. Socio-demographic characteristics of the participants had variable effects on the development of psychological symptoms of depression, anxiety and stress. Women aged between 38 - 45 years were 3 times more likely to develop depression when compared to those between 18 and 27 years (OR=3.1, 95%CI: 0.06 - 0.19, p < 0.001). Other factors significantly associated with depression among pregnant women during COVD-19-related lockdown were being para 2-4 (OR=1.9, 95%CI: 1.16-3.31, p=0.011), and occupations requiring physical contacts such as being a farmer (OR=1.9, 95%CI: 1.01-3.52, p=0.047), trader (OR=3.4, 95%CI: 0.24-0.71, p=0.001), and artisan (OR=5.1, 95%CI: 0.05-0.36, p<0.001). The factors significantly associated with lower risk of depression among the participants were being aged 18 - 27 years (OR=0.6, 95%CI: 0.37-0.9, p=0.016), para 1 (OR=0.5, 95%CI: 0.31-0.94, p=0.028), divorced (OR=0.2, 95%CI: 0.05-0.93, p=0.04) and civil servant (OR=0.5, 95%CI: 2.23-13.6, p<0.001).

**Table 4:** Multivariable logistic regression analysis showing the predictors of depression, anxiety and stress among the participants

| Variable | Depression |  | Anxiety |  | Stress |  |
| --- | --- | --- | --- | --- | --- | --- |
|  | OR(95%CI) | P value | OR(95%CI) | P value | OR(95%CI) | P value |
| Age, yr |  |  |  |  |  |  |
| 18 - 27 | 1.00 |  | 1.00 |  | 1.00 |  |
| 28 - 37 | 0.6 (0.37-0.9) | 0.016 | 1.0(0.64-1.61) | 0.946 | 1.5(0.95-2.22) | 0.081 |
| 38 - 45 | 3.1(0.06-0.19) | <0.001 | 2.2(0.10-0.30) | <0.001 | 0.9(0.51-1.44) | 0.559 |
| Parity |  |  |  |  |  |  |
| 0 | 1.00 |  | 1.00 |  | 1.00 |  |
| 1 | 0.5(0.31-0.94) | 0.028 | 0.7(0.39-1.27) | 0.245 | 1.5(0.31-2.81) | 0.501 |
| 2 - 4 | 1.9(1.16-3.31) | 0.011 | 1.8(0.98-3.14) | 0.055 | 0.4(0.79-2.37) | 0.051 |
| ≥ 5 | 0.7(0.38-1.21) | 0.185 | 4.1(0.07-0.26) | <0.001 | 5.9(2.99-11.9) | <0.001 |
| Marital status |  |  |  |  |  |  |
| Single | 1.00 |  | 1.00 |  | 1.00 |  |
| Married | 0.5(0.17-1.23) | 0.112 | 0.8(0.34-1.91) | 0.622 | 0.2(0.08-0.52) | <0.001 |
| Divorced | 0.2(0.05-0.93) | 0.04 | 0.5(0.14-2.18) | 0.393 | 0.5(0.12-2.15) | 0.355 |
| Widowed | 0.5(0.11-1.85) | 0.274 | 0.4(0.12-1.49) | 0.179 | 1.0(0.24-4.37) | 0.972 |
| Area of residence |  |  |  |  |  |  |
| Rural | 1.00 |  | 1.00 |  | 1.00 |  |
| Urban | 0.8(0.53-1.13) | 0.179 | 2.6(0.39-0.87) | 0.008 | 3.3(0.18-0.39) | <0.001 |
| Educational qualification |  |  |  |  |  |  |
| No formal education | 1.00 |  | 1.00 |  | 1.00 |  |
| Primary | 2.2(0.89-5.19) | 0.087 | 2.3(0.95-5.41) | 0.065 | 0.7(0.29-1.94) | 0.547 |
| Secondary | 0.9(0.42-1.88) | 0.758 | 1.6(0.73-3.32) | 0.248 | 0.2(0.09-0.47) | 0.421 |
| Tertiary | 1.3(0.6-2.89) | 0.484 | 3.8(1.67-8.43) | 0.001 | 0.1(0.08-3.46) | 0.301 |
| Occupation |  |  |  |  |  |  |
| House wife | 1.00 |  | 1.00 |  | 1.00 |  |
| Farmer | 1.9(1.01-3.52) | 0.047 | 0.9(0.43-1.73) | 0.689 | 0.6(0.34-1.11) | 0.108 |
| Trader | 3.4(0.24-0.71) | 0.001 | 4.2(0.12-0.39) | <0.001 | 0.1(0.07-4.21) | 0.089 |
| Artisan | 5.1(0.05-0.36) | <0.001 | 0.3(0.11-1.59) | 0.061 | 1.4(0.64-3.43) | 0.355 |
| Civil servant | 0.5(2.23-13.6) | <0.001 | 1.7(0.71-4.13) | 0.229 | 1.5(0.75-3.04) | 0.246 |
| Semester of gestation |  |  |  |  |  |  |
| 1<sup>st</sup> trimester | 1.00 |  | 1.00 |  | 1.00 |  |
| 2<sup>nd</sup> trimester | 0.7(0.42-1.17) | 0.173 | 1.5(0.83-2.69) | 0.179 | 1.2(0.85-5.54) | 0.361 |
| 3<sup>rd</sup> Trimester | 1.1(0.65-1.88) | 0.710 | 0.2(0.09-1.29) | 0.074 | 1.9(1.05-3.29) | 0.034 |

Age 38 - 45 years (OR=2.2, 95%CI: 0.1-0.3, P<0.001), para 5 and above (OR=4.1, 95%CI: 0.07-0.26, p<0.001), urban residence (OR=2.6, 95%CI: 0.39-0.87, p=0.008), tertiary education (OR=3.8, 95%CI: 0.12-0.39, p=0.001) and being a trader (OR=4.2, 95%CI: 0.12-0.39, p<0.001) were variables significantly associated with anxiety among the study cohorts.

Higher parity (≥5), urban residence and being in 3^rd^ trimester gestation were significantly associated with risk of development of stress symptoms. Grandmultiparous women were nearly 6 times more likely to report stress symptoms when compared to nulliparous participants (OR=5.9, 95%CI: 2.99-11.9, p<0.001). Also participants who lived in urban centers were 3 times at higher risk of reporting stress symptoms when compared with those living in rural areas (OR=3.3, 95%CI: 0.18-0.39, p<0.001). Stress was about twice higher in women in their 3^rd^ trimester of pregnancy when compared to those in the 1^st^ trimester (OR=1.9, 95%CI: 1.05-3.29, p=0.034).

## DISCUSSION

Although pregnancy is a time of joy for most women, some women experience a range of negative emotions during pregnancy leading to anxiety and depression.^13^ Maternal mental health problems are associated with short-term and long-term risks for the affected mothers’ overall health and functioning, as well as their children’s physical, cognitive and psychological development.^14^ Conditions such as extreme stress, emergency and conflict situations, and natural disasters can inflate the risks of perinatal mental health morbidity.^15^ Therefore, it is plausible that pregnant women are vulnerable to mental ill-health during the COVID-19 pandemic.

Several studies on COVID-19 and pregnancy have been published recently, but few studies have evaluated the impact of this pandemic on maternal mental health particularly in low-resource setting. In this study, we evaluated the prevalence of depression, anxiety and stress among pregnant women during COVID-19-related lockdown.

We found high level of depression (45.2%) among the study participants. Pregnant women who were between the age of 38 - 45 years, para 2-4 and were in occupation requiring physical contacts such trading were more likely to be affected. Over one-third (37.5%) of pregnant women reported COVID-19-related anxiety. Similar findings were reported by Taubman-Ben-Ari et al. in Israel where a significant number of pregnant women reported COVID-19-related anxiety, with the greatest concern due to being in public places or taking public transport. This was followed by anxieties about the health of others, either the foetus or family members, and then the possibility of the pregnant woman herself being infected and anxiety about the delivery. Whereas the same anxieties are shared by pregnant women characterised by diverse sociodemographic variables, the lockdown had resulted in low economic activity and difficulty assessing transportation in the event of emergency. These conditions may make a pregnant woman feel less secure and therefore feel more anxious and distressed in the face of the current crisis.

Similarly, over half (56.8%) of the participants reported stress symptoms. High parity (≥5), urban residence and being in 3rd trimester gestation were significantly associated with risk of development of stress symptoms. This is not surprising as women in these group need social support to lightened the burden of pregnancy. Social distancing has resulted in inadequacy of social supports for many pregnant women as they are separated from their loved ones, whom they rely on for support during this critical phase. While the social support helps pregnant women to relieve stress, inadequate social support is one of the significant risk factors for extreme stress and depression among pregnant women.

The long-term mental health outcome of COVID-19 is currently not known and it might take months to be completely evident. To provide adequate support to pregnant women in the developing countries which has minimum mental health services available, there is a need to integrate screening for anxiety, stress and depression in existing antenatal care programs. A detailed mental health crisis program should be developed by introducing innovative strategies like tele-consultation services to give psychological assistance to pregnant mother to deal with secondary mental health challenges related with COVID-19. There is need for more research to assess the impact of this pandemic on pregnant women and their babies particularly in developing countries, where mental health care is poorly developed. Culture sensitive intervention programs should be put in place and also be taught to healthcare workers such as nurses and midwifes for early intervention for pregnant women suffering from COVID-19-related psychological distress.

This study has a limitation. It is a single centre study that cannot be considered representative of the population of pregnant women in Nigeria during the lockdown period. A multicentre study would have improved the generalization of the study findings. Despite this limitation, this is a timely study and reflects the psychological distress of pregnant women in the face of the spread of COVID-19.

In conclusion, a significant proportion of pregnant women had psychological symptoms of depression, anxiety and stress during COVID-19-related lockdown in Nigeria. There is a need to pay special attention to vulnerable populations, such as pregnant women, so as to prevent long term adverse psychological outcome that may result from COVID-19 pandemic.

## Data Availability

Data is available upon request

## Conflict of interest

Authors declare no conflict of interest

## Acknowledgement

None

## Notes

### Competing Interest Statement

The authors have declared no competing interest.

### Author Declarations

Research and Ethics Committee of Alex Ekwueme Federal University Teaching Hospital,Abakaliki,Nigeria

## REFERENCES

1. Nwafor JI, Aniukwu JK, Anozie BO, Ikeotuonye AC, Okedo-Alex IN. Pregnant women’s knowledge and practice of preventive measures against COVID-19 in a low-resource African setting. Int J Gynaecol Obstet. 2020; 150(1): 121–123. doi:10.1002/ijgo.13186.

2. Nigeria Centre for Disease Control. An update of COVID-19 outbreak in Nigeria. 2020.https://ncdc.gov.ng/diseases/sitreps/?-cat=14&name=An%20update%20of%20COVID-19%20outbreak%20in%20Nigeria. Accessed August 24, 2020.

3. Kajdy A, Feduniw S, Ajdacka U, Modzelewski J, Baranowska B, Sys D, et al. Risk factors for anxiety and depression among pregnant women during the COVID-19 pandemic: a web-based cross-sectional survey. Medicine 2020;99:30(e21279).

4. Verbeek T, Arjadi R, Vendrik JJ. Anxiety and depression during pregnancy in Central America: a cross-sectional study among pregnant women in the developing country Nicaragua. BMC Psychiatry 2015;15:292.

5. Lee CH, Huang N, Chang HJ, Hsu YJ, Wang MC, Chou YJ. The Immediate Effects of the Severe Acute Respiratory Syndrome (SARS) Epidemic on Childbirth in Taiwan. BMC Public Health. 2005; 445:(51):30 doi:10.1186/1471-2458-5-30.

6. Taubman-Ben-Ari O, Chasson M, Sharkia SA Weiss E. Distress and anxiety associated with COVID-19 among Jewish and Arab pregnant women in Israel, J Reprod Infant Psych 2020; 38:(3) 340-348. DOI:10.1080/02646838.2020.1786037.

7. Tomfohr-Madsen L, Cameron EE, Dunkel Schetter C, Campbell T, O’Beirne M, Letourneau N. Pregnancy anxiety and preterm birth: The moderating role of sleep. Health Psychology 2019; 38(11): 1025–1035. https://doi.org/10.1037/hea0000792

8. Yedid Sion M, Harlev A, Weintraub AY, Sergienko R., Sheiner E. Is antenatal depression associated with adverse obstetric and perinatal outcomes? J Maternal-Fetal Neonatal Medicine 2016;29(6): 863–867. https://doi.org/10.3109/14767058.2015.1023708

9. Thapa SB, Mainali A Schwank SE, Acharya G. Maternal mental health in the time of the COVID-19 pandemic. Acta Obstet Gynecol Scand. 2020;99:817–818.

10. Oliobi WC, Nwafor JI, Ikeotuonye AC, Nweke NA, Uche-Nwidagu NB, Okoye PC et al. Pattern of antenatal care among antenatal clinic attendees at Alex Ekwueme Federal University Teaching Hospital Abakaliki, Nigeria. Int J Res Med Sci 2019;7:4096-101.

11. Dean AG, Sullivan KM, Soe MM. OpenEpi: Open Source Epidemiologic Statistics for Public Health, Version. Updated 2013. www.OpenEpi.com. Accessed March 3, 2020.

12. Nwafor JI, Obi VO, Obi CN, Ibo CC, Ugoji DPC, Onwe BI, et al. Mental health outcome and perceived care needs of women treated for a miscarriage in a low-resource setting. Trop J Obstet Gynaecol 2020;37:85-94.

13. Ali NA, Feroz AS. Maternal mental health amidst the COVID-19 pandemic. Asian J Psych 2020; 54: 102261. https://doi.org/10.1016/j.ajp.2020.102261

14. Corbett GA, Milne SJ, Hehir MP, Lindow SW, O’connell, MP. Health anxiety and behavioural changes of pregnant women during the COVID-19 pandemic. Eur. J. Obstet. Gynecol. Reprod. Biol. 2020; 249, 96.

15. Ravaldia C, Wilsond A, Riccac V, Homerd C, Vannaccia A. Pregnant women voice their concerns and birth expectations during the COVID-19 pandemic in Italy. Women Birth 2020. https://doi.org/10.10167j.wombi.2020.07.002

